# Beyond buzzwords: The challenges of interdisciplinarity – *An analysis of an interdisciplinary summer school on snakebite envenoming*

**DOI:** 10.1101/2024.04.26.24306415

**Authors:** Deborah Hosemann, Jade Rae, Jörg Blessmann, Maik Damm, Ulrich Kuch, Tim Lüddecke, Benno Kreuels

**Affiliations:** Working Group Snakebite Envenoming, Department of Implementation Research, Bernhard Nocht Institute for Tropical Medicine, Hamburg, Germany; Institute for Insect Biotechnology, Justus-Liebig University Giessen, Gießen, Germany; LOEWE-Centre for Translational Biodiversity Genomics, Frankfurt am Main, Germany; Institute of Occupational, Social and Environmental Medicine, Goethe University Frankfurt, Frankfurt am Main, Germany; Department for Bioresources, Fraunhofer Institute for Molecular Biology and Applied Ecology, Gießen, Germany

## Abstract

**Background:** Interdisciplinary approaches are particularly important when it comes to complex research areas such as snakebite envenoming. To achieve the World Health Organization’s (WHO) goal of halving the number of deaths and disabilities from snakebite by 2030, researchers and experts from different fields need to work together. To promote interdisciplinarity in snakebite research and educational work, a one-week hybrid summer school was organised in September 2023 at the Bernhard Nocht Institute for Tropical Medicine, Hamburg, Germany. The week’s topics were arranged logically, from snake biology and venomics to clinical implications, new therapeutics and public policy. All lectures were held in English.

**Methodology/Principal Findings:** Attendance was recorded for in-person and online participants, transcribed into Excel, and anonymised. Data were then summarised according to the participant’s field of expertise, country of residence, and attendance at each session.

**Results:** The summer school successfully promoted interdisciplinarity, with individuals attending from a wide range of scientific fields. However, fluctuations in attendance over the week highlight some challenges in maintaining interdisciplinarity at such events. By mode of attendance, in-person participants attended more of the sessions (76.9%) than those joining in a hybrid format (50.0%) or online only (32.0%). Among those who did not attend all sessions, attendance was highest on Monday (77.6%) and Wednesday (81.3%) but decreased for individuals from all fields over the week.

**Conclusion/Significance:** For future international interdisciplinary events, we suggest hybrid events with in-person and online options to encourage more international participation, supported by travel grants. However, the online experience could be improved through online networking and interdisciplinary activities. Future events should also consider hosting events in low- and middle-income countries or satellite locations. An appeal to organizers of future events is that participant data should be collected, analysed and published for continuous improvement of such interdisciplinary events.

**Author Summary:** Promoting collaboration between researchers from different disciplines is important for improving the understanding of and ability to tackle complex research areas. However, it is unclear whether courses designed to promote interdisciplinarity are successful or whether interdisciplinarity remains merely a buzzword. To determine the success of interdisciplinary courses and identify areas for improvement, attendance and participant satisfaction during these courses must be assessed. Interdisciplinarity is particularly important for strongly interlinked fields, such as One Health topics involving humans, animals, and the environment. One of these research areas is snakebite envenoming, which poses a health threat to millions of people worldwide. We organised a snakebite summer school in September 2023 to promote interdisciplinarity in this area. The data analysed here shows patterns in participation, highlighting where interdisciplinarity was achieved and where it was lacking. Based on these findings, we recommend hybrid events that allow in-person and online attendance but suggest a range of approaches to improve the experience of online attendees, including providing access to online networking opportunities, and coordination between international organisations to allow for some in-person events in satellite locations for those who are unable to attend the main event location in person.

## Introduction

The Sustainability Development Goals set out by the United Nations in 2015 emphasise the need for collaborative and interdisciplinary efforts to address some of the complex and interlinked issues facing our world (1). Several studies have supported this approach, showing that scientific innovation is improved through collaboration between scientists from different fields (2, 3). Interdisciplinary approaches are particularly important in tackling complex research areas (1), with snakebite envenoming research standing out as a prime example (4, 5). Knowledge of the behaviour and distribution of snakes, snake venom, snakebite incidence, the social impact, community beliefs and practices, and the clinical management of snakebite need to be improved in many countries where snakebite envenoming is common. Research on snakebite envenoming is said to have “often lacked the necessary interdisciplinarity and inherent opportunities for innovation that result from collaborating across diseases, disciplines, and sectors” (6).

To achieve the World Health Organisation (WHO) goal of halving the number of snakebite deaths and disabilities before 2030 (7), researchers from different fields must work together, and researchers from the social sciences need to be more involved in snakebite envenoming research to improve our understanding of and tackle the burden of this neglected tropical disease (NTD) (5, 8).

To foster interdisciplinarity in snakebite envenoming research, a week-long hybrid summer school was organised at the Bernhard Nocht Institute for Tropical Medicine in Hamburg, Germany, in September 2023. The school brought together researchers from different backgrounds, facilitated exchange between the disciplines, and introduced these disciplines to early-career researchers working on snakebite envenoming. To assess the success of this approach and provide recommendations for facilitating interdisciplinary events, we analysed the participant data collected to monitor the in-person and online attendance of individuals from different disciplines in relation to their field.

## Methods

### Content and format of the summer school

In November 2022, an interdisciplinary network meeting on snakebite envenoming was organised by the Snakebite Envenoming working group at the Bernhard Nocht Institute for Tropical Medicine. During this one-day meeting, PhD students and other researchers of different career stages presented their work and received feedback and suggestions from renowned snakebite experts. At this meeting, the idea of an interdisciplinary summer school was born and subsequently further developed with the support of the German Alliance for Global Health Research (GLOHRA) (9).

The topics taught during the summer school were organised in a logical order, from snake biology and venomics to clinical impact, novel therapeutics and public policy. Several experts were proposed for each topic and contacted by the organisers. Each day, the summer school sessions were organised by research area: herpetology/biology (Monday), venomics (Tuesday), clinical management and community engagement (Wednesday), epidemiology and social sciences (Thursday), and an outlook on novel therapeutics and public policy (Friday). Days consisted of two 90-minute sessions on Monday and Friday and four 90-minute sessions on Tuesday, Wednesday, and Thursday. The timetable of the summer school is provided in the supporting information (S1 Appendix). All lectures were held in English language.

Slack (10) was introduced as a free online communication platform, and an invitation was extended to all participants and lecturers to facilitate online networking. The platform had five channels: 1) an introduction channel, where participants and lecturers could introduce themselves and share their contacts or professional profiles; 2) a presenters channel, where participants could post questions not answered during the sessions, which were then forwarded to the lecturers and available online; 3) a regional experiences channel, where questions on snakebite envenoming were posted to facilitate discussion on different experiences; 4) a socialising channel and; 5) a channel where documents and resources from the summer school were posted.

### Promotion and enrollment of the summer school

The event organisers and invited lecturers advertised the summer school through various channels (social media, mailing lists, etc.), and organisations in the clinical, herpetological, social sciences, and venomics fields distributed information about the summer school amongst their colleagues and students. Entry requirements were kept to a minimum, but applicants were required to submit a short paragraph demonstrating their interest in the course. Applicants could also submit a short abstract of their work to present at the summer school. Attendance, both in-person and online, was free of charge. Applicants who submitted an abstract and came from a low- or middle-income country (LMIC) could apply for a travel grant for which only limited funding was available. Five such travel grants were awarded to early career researchers and were funded by GLOHRA as part of the Global Health Academy for Innovative Training Offers.

### Data collection and management

Each participant’s field of expertise, country of residence, and attendance at each session were recorded. The fields of expertise were summarised as biology (biology, herpetology and ecology), epidemiology, medical sciences (human and veterinary medicine, clinical toxicology and other specialisations), molecular sciences (venomics, biochemistry, biotechnology and bacteriology), social sciences (health economics, public policy and health administration) and other (non-governmental organisation or funding agency representatives, computer science, and generally interested people). The countries of residence were grouped into the six WHO regions: Africa, the Americas, the Eastern Mediterranean, Europe, Southeast Asia, and the Western Pacific.

In-person attendance was recorded on printed attendance lists at least twice during the day to confirm that participants stayed for most of the day, while online attendance was recorded as the time spent in the online session on Zoom. Participants knew their attendance was being recorded as proof of attendance required to obtain the certificate of completion at the end of the summer school. The person responsible for registration compiled the in-person and online attendance data in Excel and subsequently anonymised and summarised the data for further analysis. The funder created an evaluation questionnaire distributed to all participants and lecturers who answered anonymously. This questionnaire is provided in the supporting information (S2 Appendix).

### Data analysis

Attendance each day was summarised as the total number of individuals who attended online or in-person by their field of expertise and region of residence. Online attendance was only counted for participants who were online for at least 60 minutes that day (and counted as absent otherwise). To investigate possible fluctuations in attendance, the number of participants was summarised each day for all participants and for those who were absent at least one day. The percentage of course time in attendance for online participants was calculated as the total minutes in attendance over the daily session time and was capped at 100%. The average of this value was then calculated for each day and field of expertise (which included 0 minutes online for those enrolled online but not in attendance).

### Ethical approval

A protocol for analysis of the data was submitted to the Ethics Committee of the Hamburg State Medical Association for approval. Since the data subject of the study was anonymised and can no longer be attributed to a human being, the committee approved the project as not requiring consultation with the Ethics Committee of the Hamburg Medical Association (2023-300415-WF).

## Results

The total number of participants who attended the summer school was 112. After excluding individuals who attended online for less than 60 minutes each day, the number of participants was 107. The five excluded individuals were only briefly present on one day each, three on Monday and two on Wednesday.

Overall, individuals from medical sciences accounted for the majority of participants (34.6%, 37/107) and the greatest proportion of online attendees (37.3%, 28/75) (Figure 1A). By region, attendance was highest from Europe (36.4%, 39/107), followed by Southeast Asia (20.6%, 22/107), Africa (19.6%, 21/107), and the Western Pacific (15.9%, 17/107). The majority of in-person participants came from Europe (38.5%, 10/26), followed by Africa (26.9%, 7/26) and the Western Pacific (23.0%, 6/26). Online participants were also most often from Europe (30.7%, 23/75), closely followed by Southeast Asia (26.7%, 20/75), Africa (18.7%, 14/75) and the Western Pacific (14.7%, 11/75). Individuals from the Americas only attended online (online: 5.3%, 4/75) (Figure 1B). The six participants who attended in a hybrid format were all from Europe.

**Figure 1.**
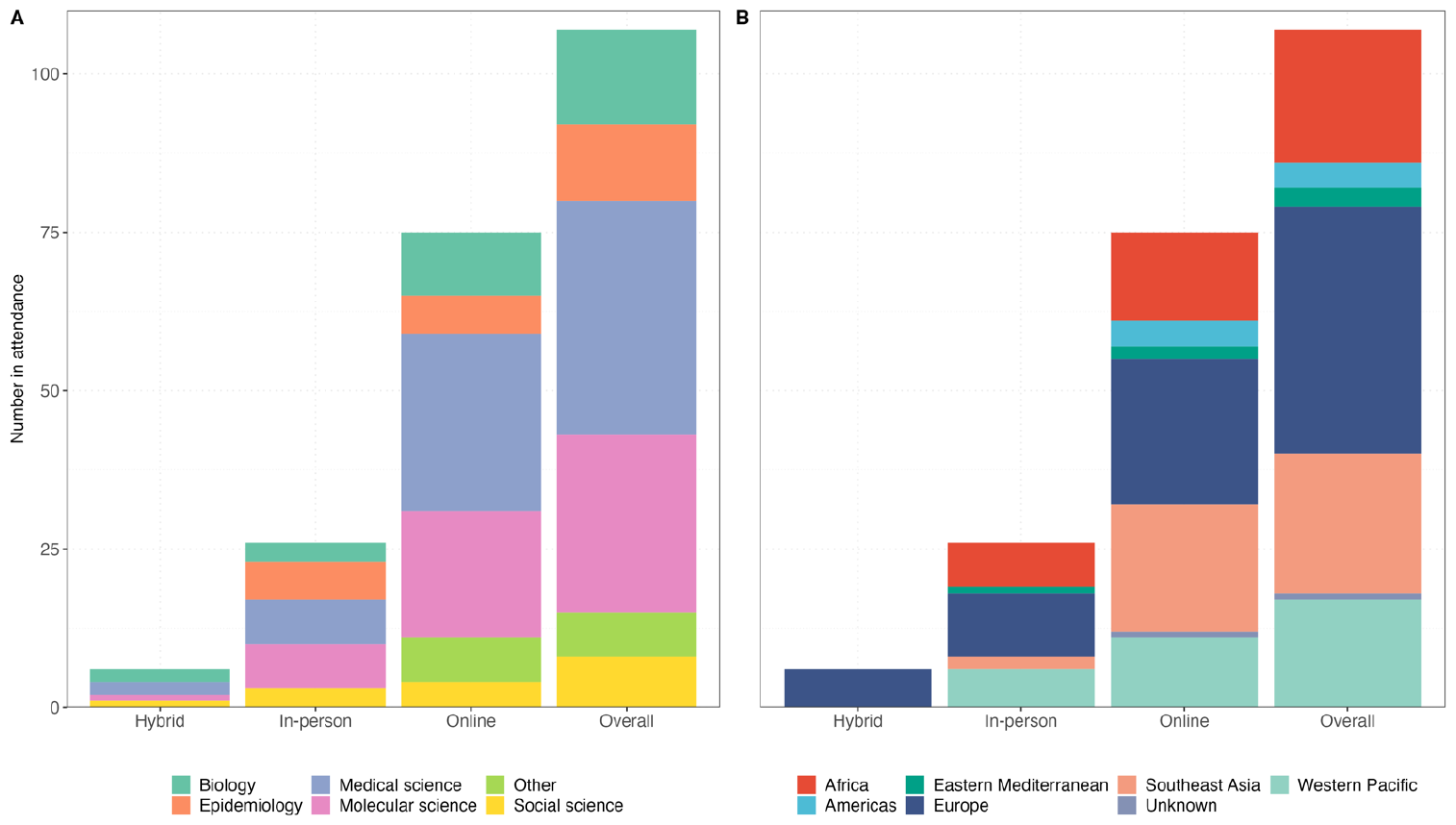
Number of participants by attendance mode and (A) field of expertise, and (B) region of residence.

Overall, 43.9% (47/107) of individuals attended all sessions, with fluctuations in attendance over the week (Figure 2A). By mode of attendance, 76.9% (20/26) of in-person participants attended all days, 50.0% (3/6) of hybrid participants attended all days, and 32.0% (24/75) of the online participants attended all days. Figure 2B shows attendance each day among those absent on at least one day – the majority of participants were present on Monday (77.6%, 83/107) and Wednesday (81.3%, 87/107), and attendance of individuals from all fields decreased over the week with the lowest attendance on Friday (51.4%, 55/107). The median number of days in attendance for individuals who did not attend on all days was 3 (25^th^ percentile—75^th^ percentile: 2 – 4 days).

**Figure 2.**
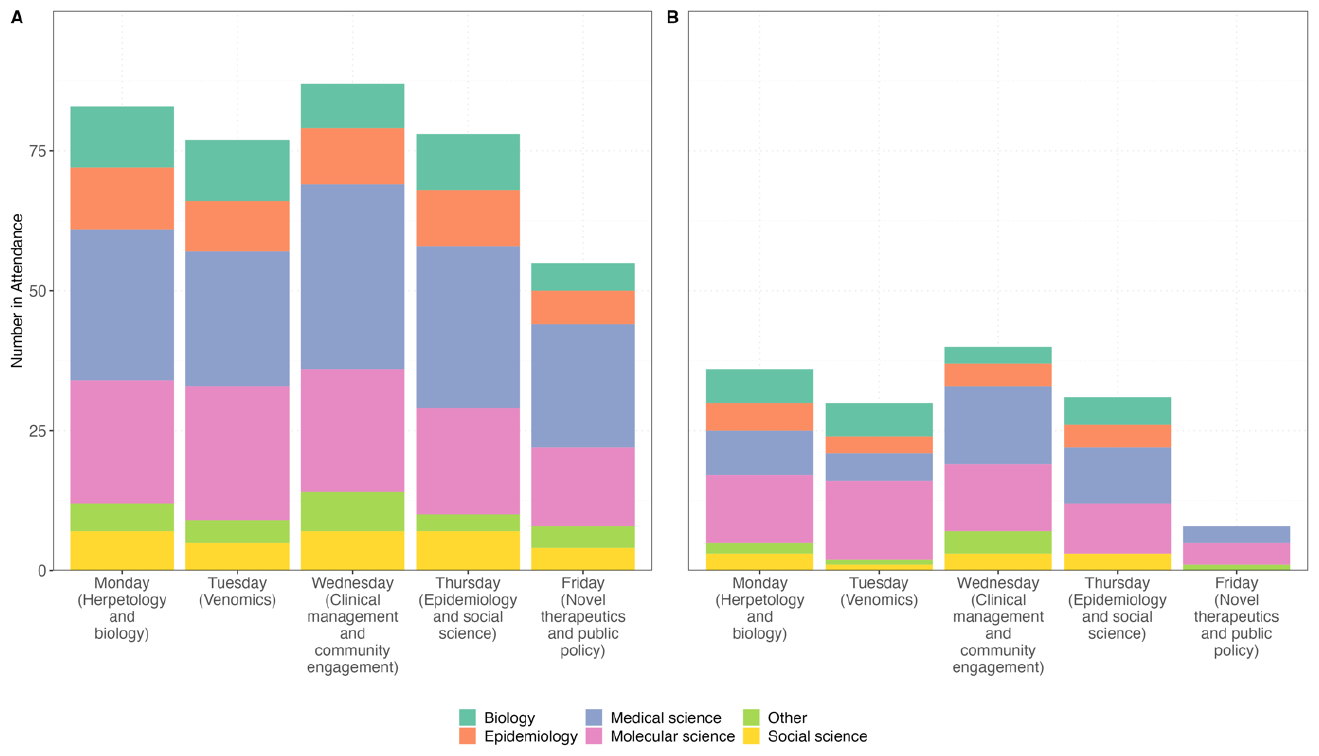
Daily attendance by field of expertise for (A) all participants and (B) participants who did not attend every day of the summer school. Online attendance each day was only considered for those who attended at least 60 minutes of course time.

Overall, the average percentage of session time spent online among online attendees was highest on Monday (64.6%, range: 49.3 – 100%) and lowest on Friday (39.6%, range: 21.9 – 54.7%). Over the week, attendance online decreased, with a slight increase on Wednesday (Figure 3), primarily among those from medical sciences, social sciences, and “other” (Figure 3).

**Figure 3:**
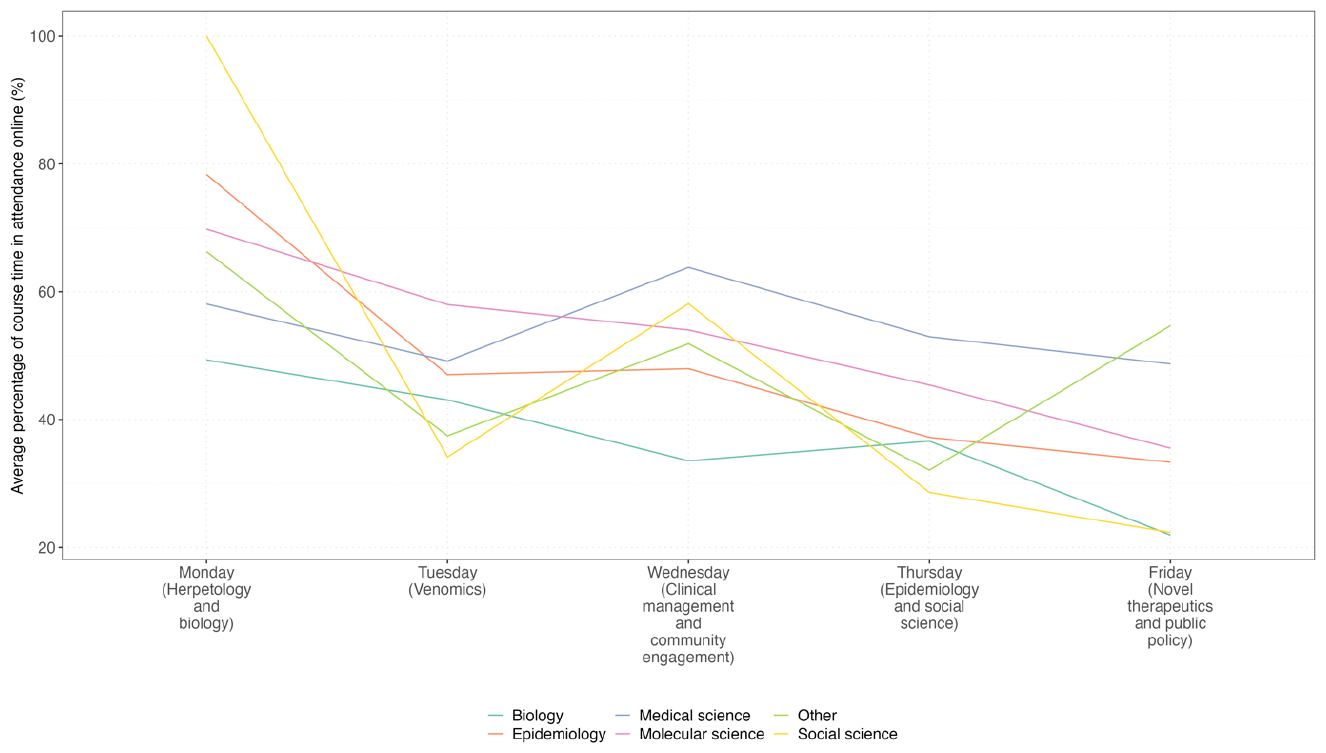
Average percentage of course time in attendance online by day and field of expertise. The percentage of time spent online for online attendees was calculated as the total number of minutes online for each discipline on each day divided by the total minutes of course time that day and was capped at 100%.

A post-training questionnaire was completed by 72 individuals, representing roughly two-thirds of all participants. From the questionnaire responses, 20.8% (12/72) of participants highlighted the importance of the interdisciplinary approach in their open field comments, and two of those also called for additional group work to build on interdisciplinarity (“*Perhaps joint group work from different disciplines would also be an option*.”). In-person attendees commented on the great networking possibilities during the course. In contrast, online attendees pointed out the lack of networking possibilities and called for more travel grants (“*I attended the summer school virtually. Unfortunately, I hardly had an opportunity to expand my network. It would be great if there were more funds for participants from low to middle-income countries*.*”*). Further, some comments confirmed that external factors played a role in variances in online attendance (*“My agenda did not allow me to attend to all the lectures* [*…*]*”*).

## Discussion

Interdisciplinary cooperation is crucial in combating NTDs. It enables holistic solutions that consider not only biomedical factors but also social determinants of health (11). Innovative strategies can be developed by integrating knowledge from different fields to reduce the burden of NTDs. This is particularly relevant for snakebite envenoming, a complex one-health challenge requiring an interdisciplinary approach for practical and effective solutions (4). To foster this approach, we organised a summer school in 2023 with lectures covering various areas of snakebite envenoming research to bring together individuals from different disciplines and backgrounds interested in this topic. Data collected on participant attendance during this course show that this approach successfully promoted internationality and interdisciplinarity, bringing together individuals from various regions and fields of expertise. However, fluctuations in attendance indicate factors that may play a role in attendance, and areas for improvement when organising future events.

### Factors influencing drop-offs

Individuals who attended the summer school in person participated in almost all sessions except on Thursday and Friday. This decrease was also observed for online participants on Friday and could be due to several factors, including the “Friday effect”, which refers to higher absenteeism on Fridays. A similar trend has been observed in other analyses of regular school attendance and, therefore, does not necessarily correspond to the content or topics of the lectures on that day (12). Other factors could include 1) student factors, including a lack of interest in the content, lack of academic background or skills necessary to follow the subject, or lack of motivation; 2) environmental factors, including other work, family, or study commitments, or other external factors such as internet or power issues and time zone differences, and 3) course factors, including lectures not being engaging enough or the lack of institutional support (13).

Individuals who attended the summer school in person participated in more sessions than those who attended online, highlighting one of the main challenges of online courses. Most individuals who attended in person had to travel from outside of Hamburg (the location of the summer school), so the cost of travel, borne by themselves, their institute, or funded by a travel grant, could have been an incentive to attend more of the sessions. In contrast, online participants likely had other obligations during the week and could therefore not attend all sessions, as reflected in the evaluation comments and commonly reported in online courses (13, 14). Further, the absence of in-person attendees is more easily noticed, so their higher attendance rates could be associated with a sense of responsibility to themselves, their institutions, the organiser, or the funder.

Another obstacle to successfully running interdisciplinary courses is making course content interesting for individuals from a wide range of fields. The content taught on Friday, which focused on novel therapies for snakebite envenoming and public policy, could have led to lower attendance due to the existing research-policy divide (15) or because the details on new therapies presented were too far from many participants’ realities in LMICs. Content on these topics may need to be planned and advertised differently on the curriculum at future events.

### Advantages and disadvantages of in-person and online attendance

As the attendance of online participants was, on average, lower than that of in-person participants, the question arises as to whether events would be more interdisciplinary if they were only hosted for in-person attendance. However, while this analysis shows that attendance was more consistent among in-person participants, a hybrid model (with sessions that have both online and in-person attendants) improves the internationality of such training sessions, which can be an important factor in fostering interdisciplinarity. Hosting sessions in a hybrid mode has several advantages, including reducing costs incurred by the host organisation as well as participants and not requiring participants to travel to the course, which decreases the environmental impact of the event by reducing long-distance travel (16). These costs are often a barrier, particularly for participants from LMICs, especially since such events are often hosted in high-income countries (17). Therefore, the possibility of online attendance and other facilitators like travel grants increases the accessibility of these courses (18). Only five such travel grants were available in our case, although the demand was much higher. Many applicants who were not awarded a travel grant participated online instead. However, it should be noted that this is not a long-term solution to achieve “conference equity”, and other solutions, such as hosting such events in LMICs, should be considered by organisers (17). While online participation has many advantages, it also has several disadvantages. In-person attendance allows for networking outside of course time, promoting ongoing and new collaboration (18).

While Slack was introduced as a free online communication platform, and an invitation was extended to all participants and lecturers to facilitate online networking, such a platform cannot replace face-to-face dialogue and connections made during in-person attendance, as noted in other online conferences (18). Future events should look for better solutions so online participants can network better when in-person attendance is not possible. In addition, it is essential to look for ways to bridge the gap between online and in-person participants. There is often a discrepancy between online participants, who mainly interact with their online peers, and in-person participants, who mainly interact with those physically present. This challenge must be addressed to ensure an interdisciplinary and engaging event for all. To improve this, additional sessions for interdisciplinary group work or mandatory sessions for networking between online and in-person participants and speakers might be arranged.

Our analysis of interdisciplinarity and attendance is limited by the fact that we could only measure interdisciplinarity in terms of quantity, not quality. We could not determine whether participants were fully engaged while they attended in-person. Similarly, since no video was required, we could not determine whether online participants were merely logged in via their computer but not listening, or actively participating in online sessions. A more detailed, targeted collection of attendance data and post-training evaluation data at future interdisciplinary (teaching) events is needed to further identify gaps in attendance and ways to promote interdisciplinarity.

Based on the evaluation of this summer school, we have several recommendations for future interdisciplinary events. If the aim is to promote international attendance, we recommend hybrid events allowing in-person and online attendance. This should be supplemented by travel grants that are sufficient in number and volume, and awarded to participants from different disciplines and countries. To improve the experience of online participants, future events could also include greater involvement of individual online participants such as through guided, obligatory networking sessions and interdisciplinary group work for in-person and online participants. If courses are organised between international institutions, in-person attendance at satellite locations could also bridge the gap in networking opportunities for individuals who cannot travel to the main location. For future interdisciplinary events, attendance data should be collected, analysed and published to identify areas of improvement and thus improve the delivery of genuine interdisciplinary events.

## Data Availability

The underlying data is available on request.

## Acknowledgements

We want to thank all lecturers and participants for being part of this summer school.

## Supporting information captions

**S1 Appendix. The timetable of the summer school.** The summer school ran over five days (with half days on Monday and Friday) with sessions on herpetology/biology (Monday), venomics (Tuesday), clinical management and community engagement (Wednesday), epidemiology and social sciences (Thursday), and an outlook on novel therapeutics and public policy (Friday).

**S2 Appendix. Content-specific questions of the evaluation questionnaire.** The evaluation questionnaire was provided to all summer school attendees, including the lecturers, to identify areas for improvement for future events.

